# Revisiting infradian rhythms in depressive symptoms: wearable signals identify individualised mood cycles in healthy adults

**DOI:** 10.64898/2026.07.22.26358741

**Authors:** Jodie Naim-Feil, Rachel E. Stirling, Rochelle De Silva, Mark J. Cook, Andrew Zalesky, Matthew JY Kang, Malcolm Hopwood, Philippa J. Karoly

## Abstract

Circadian biology has long been implicated in mood and depressive disorders. While longer, infradian rhythms are increasingly recognised across diverse physiological processes, their relevance to mood remains poorly understood. Using long-term wearable heart rate data, this study mapped depressive symptom severity onto person-specific multiday physiological phase. Individual-specific cycles were estimated from 413 young adult participants (Notre Dame NetHealth project) alongside up to four repeated Beck Depression Inventory (BDI) assessments (955 observations). Individual’s dominant infradian rhythm (2–60 days) was estimated from heart rate using wavelet analysis, and BDI scores were indexed according to whether surveys occurred during the cycle peak or trough. Mixed-effects linear modelling examined the relationship of cycle phase with mood symptoms, and whether this relationship differed by exercise level. In those with significant cycles (360 participants, 864 observations), depressive symptom severity varied across multiday physiological phase as a function of exercise level: Cycle Phase × Exercise Group interaction, *F*(1,712.21)=5.13, p=0.024. BDI scores were higher at the peak than trough in the Low-exercise group, Δ=1.69, 95% CI [0.11, 3.28], *p*=0.037, with no peak–trough difference in the Higher-exercise group. Higher daily heart rate, lower exercise, female sex, and winter season were associated with higher BDI scores. Notably, this study introduces a “when” dimension to depressive symptom variation and provides the groundwork for clinical studies testing whether wearable-derived cycles can characterise temporal patterns of symptom vulnerability and support personalised tracking of mood dynamics.

---

Major depressive disorder is among the most disabling health conditions globally, yet many patients experience persistent or recurrent symptoms despite available treatments (1,2). This persistent treatment gap suggests that important dimensions of depression vulnerability, including the temporal structure of mood dynamics, remain under-characterised. The human brain is continuously modulated by interactions between internal physiology and external influences, with mood and behaviour fluctuating over days, weeks, and months; yet systematic study of these multiday patterns remains limited. During the 20^th^ century, studies of infradian rhythms (multiday, >24 hours) or ‘mini-cycles’ in mood (3–5) and psychiatric symptoms (6–11) suggested that symptom timing could be periodic and individual-specific. However, by the late 1990s, this line of research had become much less prominent, partly reflecting a shift in psychiatry toward categorical diagnoses and shorter clinical trials, as well as the methodological challenges of capturing symptom trajectories over longer timescales (8,12,13). As a result, individual-level patterns in depressive symptom timing remain poorly characterised (14,15). Recent advances in long-term digital monitoring, ranging from intracranial devices to consumer wearable technologies, have revealed multiday fluctuations in autonomic (16–21), neural (22–27), and endocrine (28) systems. These longer rhythms are increasingly recognised as structured and biologically informative, but it remains unclear whether depressive symptoms similarly fluctuate over multiday timescales or whether symptom vulnerability aligns with an individual’s own physiological rhythm. Here, we use long-term wearable data to estimate person-specific multiday physiological phase and test whether depressive symptoms vary systematically across peak and trough phases of these rhythms.

Biological rhythms play a critical role in mood regulation and psychiatric pathophysiology (29,30). Circadian disruption is a well-established risk factor for psychiatric disturbance (31–34), associated with heightened depressive symptoms and greater relapse vulnerability (33). Circadian rhythms are entrained by external cues such as the light-dark cycle and also possess an endogenous component that can persist in the absence of environmental cues (i.e., free-running). However, circadian rhythms represent only one layer within a broader hierarchy of biological rhythms that govern brain and body function (30). Within this hierarchy, longer infradian biological rhythms (>24 hours) remain comparatively under-studied and are often framed around menstrual or seasonal processes. However, emerging evidence suggests that multiday rhythms also include intrinsic, person-specific organisation across cardiovascular, endocrine, autonomic, and neural systems (17,18,23,28). This raises the possibility that mood symptoms may also vary with longer-timescale biological rhythms.

Initial support for this interpretation comes from early psychiatric and chronobiological studies that sought to characterise mood variation over multiday timescales. Early psychiatric studies using daily ratings and spectral or autocorrelation analyses reported stable periodicities in mood symptoms ranging from 13 to 128 days in approximately one-third of hospital inpatients with affective disorders (4,6). Comparable multiday patterns were also observed in a student cohort (5,7), and chronobiological studies tracking mood in 196 healthy students similarly reported infradian rhythms in both positive and negative affect (35). Additional longitudinal work described coupling between mood symptoms and physiology over longer timescales (36), including 48-hour mood and sleep-wake cycling in manic-depressive disorder (11), as well as multiday fluctuations in blood pressure and heart rate during depressive episodes (37). Across these early studies, mood cycles were present in both male and female cohorts (4), showed increased amplitudes in clinical groups (8) and persisted despite antidepressant treatment (9,10). Together, these findings indicate that multiday mood dynamics may be person-specific (4,8), relatively stable over time (38), and present in both clinical and non-clinical populations. However, this line of research was limited by the need for intensive clinician-led symptom tracking, challenging analytic techniques, and the absence of robust tools for continuous, non-invasive monitoring in everyday life. Subsequent longitudinal studies have more commonly examined population-level rhythms, including menstrual (39,40) and seasonal (41–44) rhythms, but these approaches usually rely on group-level mean comparisons and therefore provide limited insight into individual-level variability over extended timescales (42,44).

Recent studies in neurology have renewed interest in multiday rhythms by extending findings from cardiac and autonomic signals to brain activity. By leveraging long-term intracranial electroencephalography (iEEG) these studies revealed robust, person-specific multiday rhythms of brain activity and epileptic seizures that remain stable over years (23,25,45–48). These recordings provided direct evidence of multiday rhythmicity across neural systems, with temporal features that parallel those reported in early psychiatric studies of mood periodicity. However, iEEG is invasive and rarely feasible outside severe neurological conditions, limiting translation to psychiatric and non-clinical cohorts.

To enable translation beyond invasive recordings, researchers have focused on identifying multiday rhythms non-invasively from peripheral autonomic markers, such as body temperature (19) and resting heart rate (17,18,49). Long-term wearable-derived heart rate signals have revealed prevalent multiday rhythms in healthy adults and cardiac patients, with periodicities similar to those identified in brain excitability and seizure-cycle studies in epilepsy (16–19,49). These patterns may also extend beyond autonomic physiology with cortisol, a core stress hormone, shown to vary by multiday phase, with higher levels near cycle peaks (28). Earlier longitudinal monitoring in a bipolar disorder case study also identified multiday fluctuations in blood pressure and heart rate, with amplification of an approximately weekly component during a depressive episode (37). Given the bidirectional coupling between autonomic physiology and brain regions implicated in mood regulation (28,50–53), infradian heart rate rhythms captured from wearable devices may now provide a low-burden, scalable marker for testing whether depressive symptoms vary with multiday phase. This approach treats heart-rate phase as a marker of multiday physiological state, enabling depressive symptoms and autonomic physiology to be examined as temporally aligned, rather than assuming a unidirectional causal pathway. The present study builds on this work by testing whether depressive symptom severity varies across wearable-derived multiday physiological rhythms.

Using long-term wearable data (54), alongside repeated depression symptom assessments, provides a unique opportunity for symptom severity to be examined relative to an individual’s infradian biological timekeeping. Additionally, wearables enable objective capture of physical activity and sleep patterns, which are closely linked to mood regulation and depression vulnerability. Building on the established prevalence of heart rate rhythms in healthy adults (21), and their proposed links with autonomic imbalance and stress (28), we hypothesised that depressive symptoms would be elevated at physiological defined phases of their rhythms (e.g. at the peak compared with trough). Conceptually, this tests whether tracking an underlying infradian cycle captures information about mood vulnerability that is not reducible to transient physiological changes such as elevated heart rate. By deriving individualised temporal markers from accessible, non-invasive devices, this proof-of-concept study explores a “when” dimension of digital phenotyping: not only whether symptoms are elevated, but when vulnerability may increase. This framework may provide a candidate temporal marker for future rhythm-aware monitoring and precision-timed intervention in clinical cohorts.

## Methods

Data for the present study were drawn from the NetHealth project, a publicly available longitudinal study conducted by researchers at the University of Notre Dame, Indiana, USA. The NetHealth project followed a cohort of college students (aged 17-23 years) over four years, with data collected between 2015 and 2019 (https://sites.nd.edu/nethealth). The study was supported by the National Institutes of Health (NIH), received Institutional Review Board approval, and obtained informed consent from all participants. All data were de-identified and made publicly available through the NetHealth Project website (http://sites.nd.edu/nethealth/data-2/). Participants (*n*=692) were provided with Fitbit smartwatches (either Charge HR or Charge 2 model) for continuous monitoring throughout the study period and completed demographic, behavioural, and mood surveys. The primary analysis included participants who completed depressive mood assessments, had sufficiently long, assessment-linked heart-rate data, and exhibited significant multiday rhythmicity in their heart rate, yielding 360 participants and 864 mood survey observations. For each participant, heart rate data, sex, self-reported exercise levels, sleep duration prior to survey, depressive mood symptom scores (self-reported, there was no reported clinical diagnosis in the dataset), and environmental timing variables were extracted from the dataset (sample and assessment-linked characteristics described in Table 1):

**Table 1.**
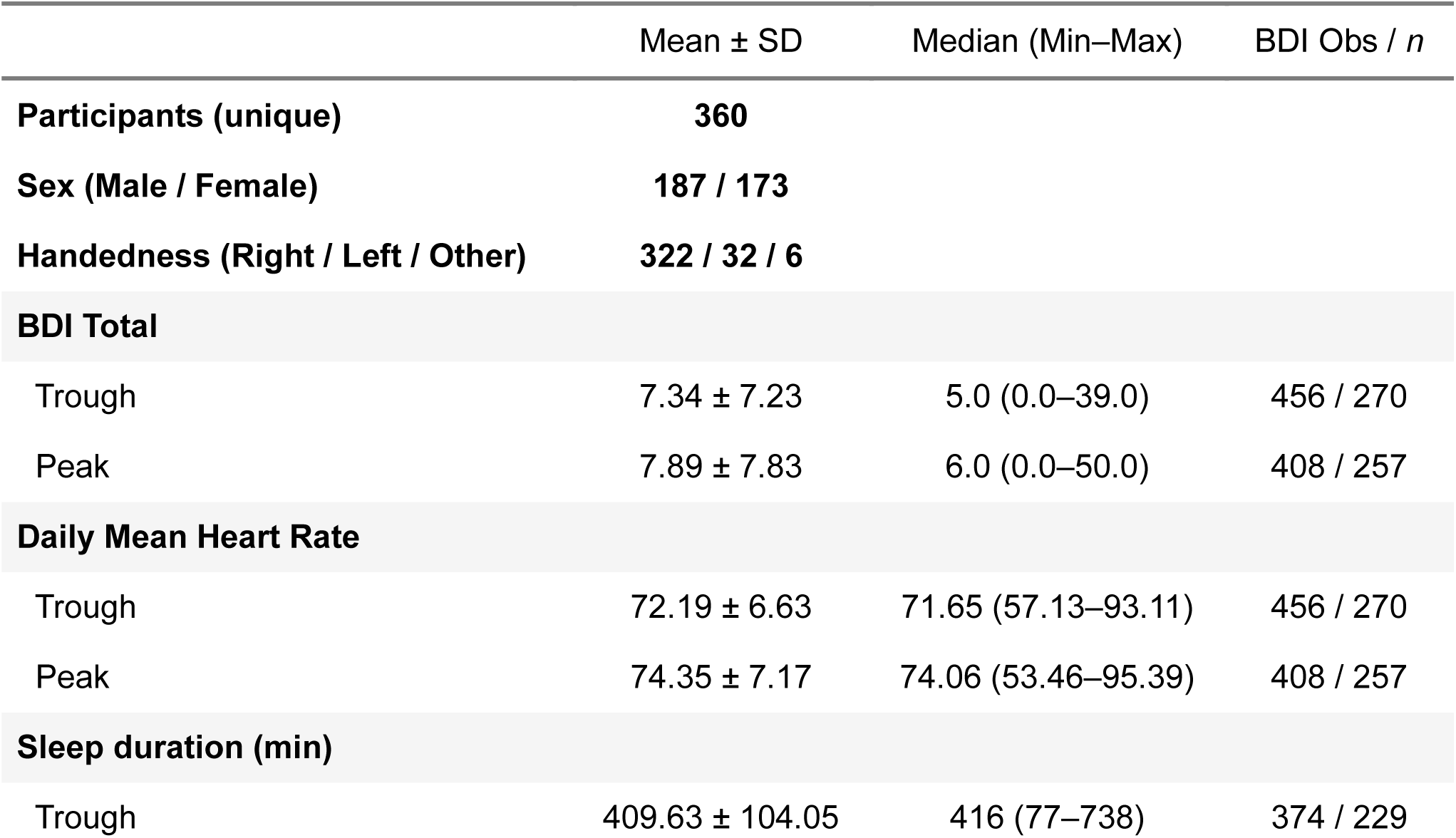
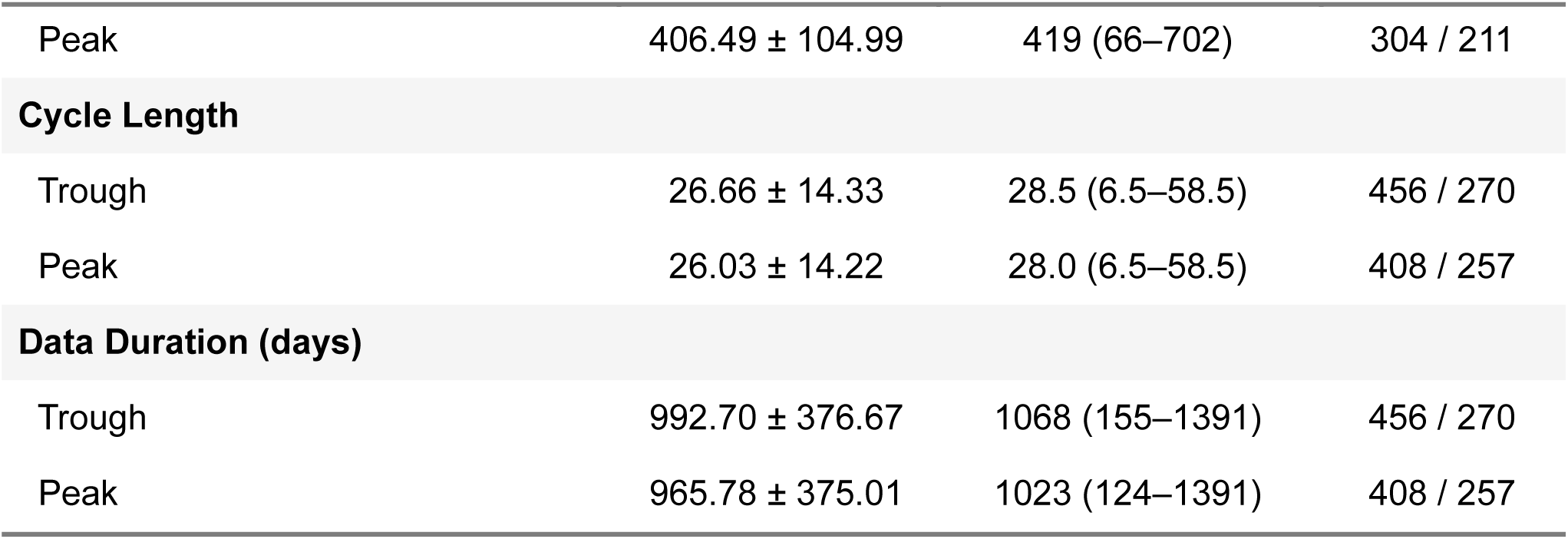
Sample and assessment-linked characteristics of the primary model. Values are shown as mean ± SD and median (min–max) unless otherwise indicated. *Obs* denotes the number of participant-session observations contributing to each phase-specific summary, and *n* denotes the number of unique participants contributing at least one observation in that phase. Participants could contribute observations to both trough and peak phases across sessions; therefore, phase-specific *N* values are not expected to sum to the total sample (*n* = 360). Variables include BDI (Beck Depression Inventory), daily mean heart rate (HR, beats per minute), minutes asleep on the night before assessment, data collection duration (days), and multiday cycle length (days). Sleep summaries are based on observations with available assessment-linked sleep data and therefore have fewer observations than the full Model A sample.

| | Mean ± SD | Median (Min–Max) | BDI Obs / *n* |
| --- | --- | --- | --- |
| **Participants (unique)** | **360** |  |  |
| **Sex (Male / Female)** | **187 / 173** |  |  |
| **Handedness (Right / Left / Other)** | **322 / 32 / 6** |  |  |
| **BDI Total** |  |  |  |
| Trough | 7.34 ± 7.23 | 5.0 (0.0–39.0) | 456 / 270 |
| Peak | 7.89 ± 7.83 | 6.0 (0.0–50.0) | 408 / 257 |
| **Daily Mean Heart Rate** |  |  |  |
| Trough | 72.19 ± 6.63 | 71.65 (57.13–93.11) | 456 / 270 |
| Peak | 74.35 ± 7.17 | 74.06 (53.46–95.39) | 408 / 257 |
| **Sleep duration (min)** |  |  |  |
| Trough | 409.63 ± 104.05 | 416 (77–738) | 374 / 229 |
| Peak | 406.49 ± 104.99 | 419 (66–702) | 304 / 211 |
| **Cycle Length** |  |  |  |
| Trough | 26.66 ± 14.33 | 28.5 (6.5–58.5) | 456 / 270 |
| Peak | 26.03 ± 14.22 | 28.0 (6.5–58.5) | 408 / 257 |
| **Data Duration (days)** |  |  |  |
| Trough | 992.70 ± 376.67 | 1068 (155–1391) | 456 / 270 |
| Peak | 965.78 ± 375.01 | 1023 (124–1391) | 408 / 257 |

### Heart rate

Fitbit wristbands were worn by all participants, capturing minute-by-minute heart rate data. For each participant, the best available segment of data was included for analysis, defined as the longest continuous segment spanning at least three months with over 70% adherence (i.e., <30% missing data). Datasets which did not meet these criteria and those with daily heart rate data values missing at the time of the survey assessment were excluded from further analysis. This data formed the basis for the extraction of multiday cycle features (described in *Heart rate cycle data extraction*).

### Depressive mood symptoms

The Beck Depression Inventory (BDI) (55): The BDI is a 21-item self-administered scale designed to assess cognitive and somatic aspects of depressive symptoms. The BDI has strong psychometric performance including high internal consistency and moderate-to-high test–retest reliability. It is a well-stablished and validated measure of depressive symptom severity (56) and administered across both clinical and non-clinical cohorts (57). Participants are asked to select the response that best describes how they have felt over the past two weeks. Depending on when participants joined the longitudinal study, they could complete the BDI assessment at up to four different timepoints (Winter 2016, Summer 2016, Spring 2017 and Spring 2018). No assessments were conducted during the Autumn season.

### Exercise

At each testing session, participants reported their typical exercise frequency over the past year using a five-point scale: not at all, less than 1–2 times per month, 1–2 times per month, 1–2 times per week, or three times per week or more. For the present analyses, responses of “not at all” or “less than 1–2 times per month” were classified as low exercise, whereas all higher-frequency responses were classified as higher exercise. Self-reported exercise level (low or higher) was assessed at each testing session, as such exercise group was treated as a time-varying behavioural variable, and participants could move between low and higher exercise categories across testing windows. Given its established association with depressive symptoms, exercise level was included in the primary model to test whether phase-related differences in symptom severity varied according to exercise.

### Sleep

Sleep duration was indexed as minutes asleep during the night prior to each BDI assessment. Sleep data was restricted to participants with sufficient repeated sleep-linked observations to estimate within-person sleep deviations, yielding n = 129 participants and 393 observations. Given the smaller subset with sufficient repeated sleep-linked observations, sleep was examined as a sensitivity analysis rather than included in the primary model. Specifically, sleep was examined in a sensitivity analysis to test whether night-before sleep accounted for, or moderated, phase-linked variation in depressive symptoms. To distinguish acute sleep variation from habitual sleep differences, sleep duration was decomposed into within-person and between-person components. For each participant, mean sleep duration from night-before was calculated across available assessment-linked sleep observations. The within-person sleep term reflected each observation’s deviation from that participant’s own mean sleep duration, whereas the between-person sleep term reflected each participant’s mean sleep duration relative to the overall cohort mean.

### Environmental effects

Environmental timing variables were examined in exploratory analyses to assess whether depressive symptom severity varied by season or day of assessment. Assessment dates were used to derive Northern Hemisphere meteorological season, consistent with the USA-based cohort, and day of week. Seasons were classified as winter, spring and summer (autumn was not assessed) using standard Northern Hemisphere month groupings.

### Heart rate cycle data extraction

Heart rate data were downloaded from the NetHealth platform, containing timestamped daily average heart rate (in beats per minute) and the corresponding daily standard deviation values (derived from minute-by-minute heart rate). Analyses were conducted at the daily level, with the survey assessment linked to the corresponding daily heart-rate value. To explore infradian rhythms in heart rate, we derived an autonomic proxy for daily resting heart rate by subtracting the daily standard deviation from the daily mean heart rate values. This proxy was intended to capture the lower bound of daily heart rate fluctuations and provide a more stable signal for rhythms extraction; it should not be interpreted as clinically measured resting heart rate.

### Wavelet transform and cycle detection

To identify multiday rhythmicity in heart rate, a continuous Morlet wavelet transform was applied to the heart rate signal. The Morlet wavelet is an established approach for detecting infradian rhythms (18,21,47), appropriate for detecting non-stationary, quasi-periodic patterns in physiological data (18). Candidate cycle periods were tested from 2 to 60 days in 12-hour increments, and peaks in the global wavelet power spectrum identified potential heart rate cycle periods for participants with sufficient recording duration (all data duration > 120 days, as presented in Table 1). The global wavelet power spectrum represents wavelet power averaged across time for each candidate cycle period.

To assess the statistical significance of detected peaks, a nonparametric permutation test was performed. Each participant’s signal was randomly shuffled 10,000 times to disrupt the temporal structure, and the wavelet transform was applied to each shuffled dataset. For each shuffled dataset, wavelet power was averaged across time to generate a global wavelet power estimate at each candidate cycle period. These permutation-derived global wavelet power values were then used to produce a null distribution for each cycle period. The 99th percentile of this null distribution was chosen as the significance threshold. Observed global wavelet power peaks in the original signal exceeding this threshold were considered statistically significant. When multiple significant peaks were identified, the most prominent cycle was selected using a peak prominence algorithm, which identifies the peak with the strongest power relative to surrounding values. Only datasets with significant multiday cycles were included for the primary analysis as cycle phase is most interpretable when derived from an identifiable physiological rhythm rather than weak or non-significant periodic structure.

### Cycle extraction from the heart rate signal

Following cycle detection, a second-order Butterworth bandpass filter was applied to isolate the dominant cycle. Cut-off frequencies were set at the upper and lower bounds of the full-width-half-maximum of the peak observed in the wavelet periodogram, to account for natural variations in biological cycle period. A schematic of the data extraction process and example data is presented in Figure 1.

**Figure 1.**
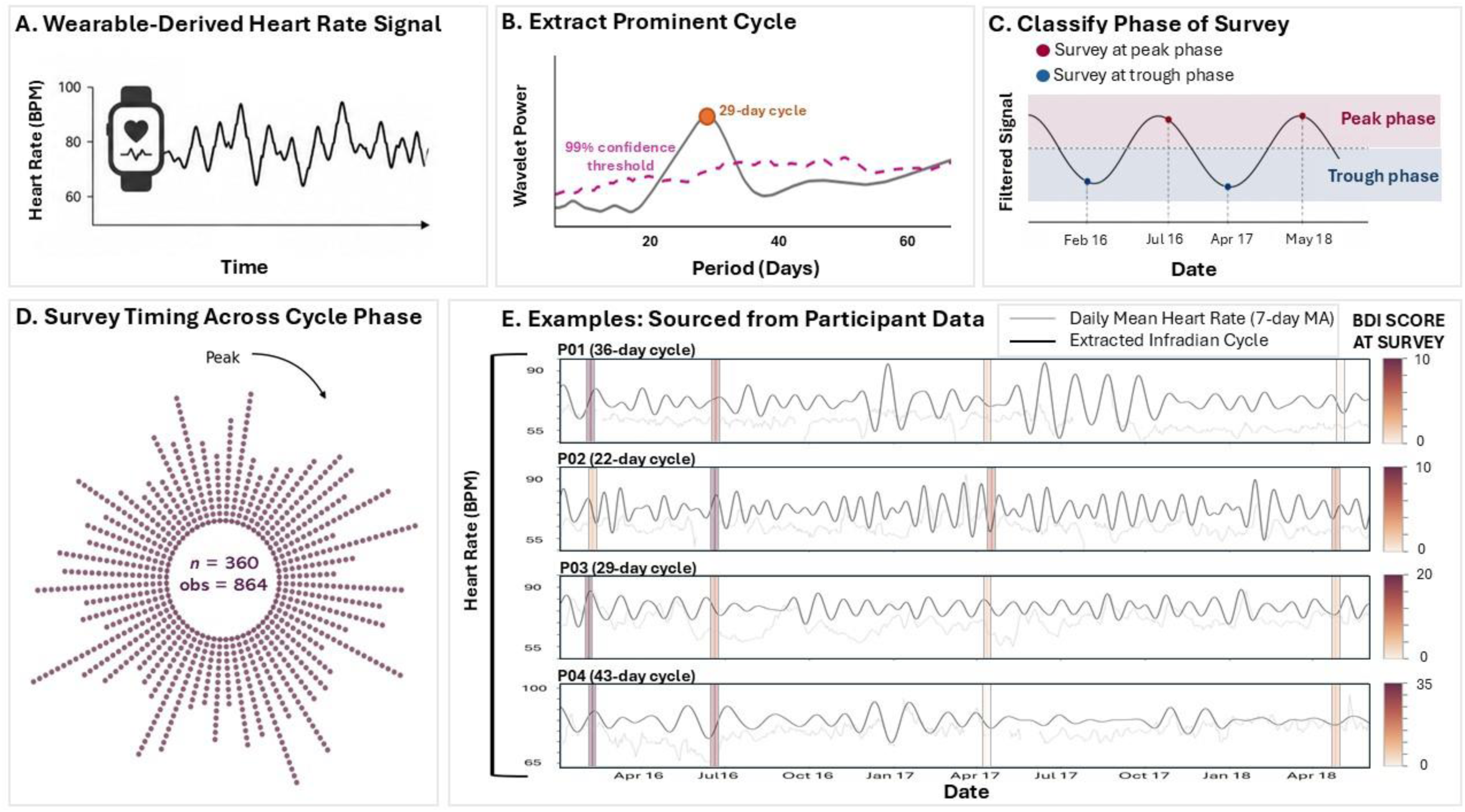
Wearable-derived pipeline for estimating person-specific multiday heart-rate phase and modelling depressive symptoms across the cycle. **1A. Wearable-derived heart rate signal:** Daily heart rate data were extracted from smartwatch data (daily mean HR derived from minute-by-minute HR) and pre-processed by selecting the longest continuous segment more than 3 months with greater than 70% adherence for each participant. A resting-HR proxy (daily mean minus 1 standard deviation) was computed to provide a more stable signal. **1B. Extract prominent cycle**: A Morlet wavelet transform was used to identify statistically significant peaks, defined as periods whose power exceeded the 99% confidence threshold. The dominant multiday peak was selected based on peak prominence, and participants without a significant multiday peak were excluded from the primary analysis. **1C. Classify phase of survey**. The prominent cycle was reconstructed using a 2^nd^ order Butterworth bandpass filter, and each survey observation classified according to its timing relative to the participant’s reconstructed infradian heart rate phase, labelled as Peak or Trough. **1D. Survey timing across phase**: Survey observations were distributed across the multiday cycle phase; each dot represents one survey observation, with 864 observations from 360 participants. Radial position is used for visualisation only and does not indicate symptom severity. **1E. Examples: Sourced from participant data**: Participant-level time series show longitudinal daily mean heart rate, reconstructed multiday heart-rate cycles, and survey timing during the primary survey testing window (2016-2018). Each row shows one participant, labelled with their estimated dominant cycle length (P1: 36 days; P2: 22 days; P3: 29 days; P4: 43 days). Grey traces indicate daily mean heart rate smoothed with a 7-day moving average, black traces indicate the reconstructed multiday heart rate cycle, and vertical colour-coded survey markers indicate survey observations, coloured according to BDI score at the time of survey.

### Identifying Cycle Phase

To assess phase-dependent effects, the filtered signal was analysed to identify whether each survey assessment occurred closer to the peak or trough of an individual’s multiday cycle. Values above the 50th percentile of the filtered signal were labelled as ‘Peak’ and values below were labelled as ‘Trough’ of the cycle. A phase label (‘Peak’ or ‘Trough’) was assigned to each survey date based on an individual’s multiday heart rate cycle. Multiday cycle phase was derived from the filtered resting-heart-rate proxy, whereas daily mean heart rate on the survey date was included separately in the statistical model as a day-level autonomic covariate. Three parameters were extracted from the heart rate data; 1. Cycle Phase, whether the survey assessment occurred near the peak or trough of the participant’s multiday cycle; 2. Daily Mean Heart Rate, the raw mean heart rate value corresponding to the survey assessment date; and 3. Cycle Length, the period in days of the participant’s dominant significant multiday cycle, ranging between 2 – 60 days.

### Data analysis

Linear mixed-effects models were applied to test whether multiday heart rate cycle phase was associated with fluctuations in depressive symptoms in non-clinical young adult cohort. Analyses were conducted in R (RStudio v2025.05.0) using *lme4* and *lmerTest*. The primary outcome was depressive symptom severity (BDI score), assessed repeatedly across sessions within individuals. All models included a random intercept for participant (Participant) to account for within-person dependence of repeated observations. Exercise was specified as the primary behavioural moderator of interest, given its established relevance to mood regulation (58,59) and its suitability for classification into interpretable activity categories, defined here as low and higher exercise. Fixed effects included cycle phase (Trough or Peak) defined by multiday cycle phase at the time of assessment; exercise level (Low vs Higher) defined from self-report exercise levels at each assessment (time-varying); and the Cycle Phase X Exercise interaction. Sex (Female vs Male) was included as a time-invariant covariate. Daily mean heart rate was included as a covariate to account for established associations between heart rate and symptom severity. Daily mean heart rate was transformed by grand-mean centring across eligible observations in the analysis sample (HR_c_), such that coefficients reflect change in BDI score per 1-Beat-per-minute (1-BPM) difference relative to the sample mean.

The primary mixed-effects model (Model A) was specified as:

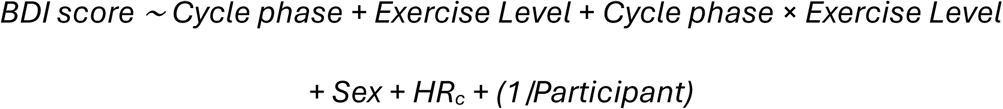

Models were estimated using restricted maximum likelihood (REML). Fixed-effect inference used Type III tests with Satterthwaite degrees-of-freedom approximations implemented in *lmerTest*. Given the Cycle Phase X Exercise Level interaction, cycle-phase effects were interpreted using planned contrasts estimated from marginal means (*emmeans*); degrees of freedom were estimated using the method available for each model. Statistical significance was defined as *p* < 0.05, two-tailed.

### Robustness across phase-sampling densities (Models A-C)

To assess robustness of the primary findings, the same fixed-effects structure was repeated across three nested samples: **Model A**: All eligible observations, **Model B,** participants with ≥ 1 observation in both Peak and Trough, and **Model C**: participants with ≥ 2 observations per phase. This approach addresses the possibility that sparse phase sampling could inflate or obscure phase-related effects, while preserving the interpretability of the fixed-effect structure across samples. Model A was specified as the primary inferential model, with Models B and C treated as sensitivity analyses reflecting increasingly stringent requirements for within-person phase coverage. Accordingly, inference focused on the pre-specified Cycle Phase × Exercise Level interaction in Model A, while sensitivity models were interpreted descriptively rather than as independent confirmatory tests.

### Sensitivity analysis including non-significant cycles

The primary analyses were restricted to participants with significant multiday heart-rate cycles, reflecting the study aim of testing whether depressive symptoms vary by the phase of an identifiable physiological rhythm. However, the permutation procedure used to define significant peaks disrupts both rhythmic structure and autocorrelation and may therefore provide a liberal test of rhythmicity. As a sensitivity analysis, the primary model (Model A) was then repeated in a broader sample that included observations from both significant and non-significant cycles (*n* = 413 participants, obs *= 955)*. This analysis tested whether the primary Cycle Phase × Exercise Level effect was reduced in the broader sample, when cycles not reaching the significance threshold were included. Cycle significance status was also included as an additional predictor in a separate sensitivity model to assess whether the primary interaction was dependent on the permutation-based significance criterion.

### Additional candidate predictors and sensitivity analyses

Sleep moderation was tested by adding a Cycle phase × sleep within-person interaction and comparing the models with and without this term using likelihood ratio tests (maximum likelihood). In the sleep sample (*n* = 129 participants; obs = 393) adding Cycle Phase × within-person sleep term did not improve model fit (likelihood ratio test χ²(1) = 0.0365, *p* = 0.8486), and neither within-person nor between-person were significant (both *p* > 0.55). Likewise, adding multiday cycle length as a covariate did not improve model fit for BDI (χ² (1) = 0.90, *p* = 0.34), and the cycle-length term was non-significant (*p* = 0.35). These variables were therefore excluded from the primary models to maintain model parsimony.

### Environmental and calendar effects

To contextualise variability in depressive symptoms across the study period, assessment dates were used to derive USA meteorological season (Northern Hemisphere) and day of week for observations from participants in Model A. Environmental effects were first examined in separate random-intercept models:

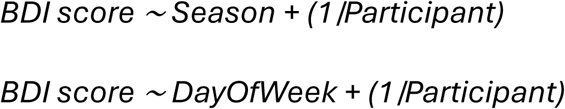

To assess whether environmental timing altered the primary findings, Season and DayOfWeek were then added as covariates to the primary mixed-effects model. Interaction terms (Cycle phase × Season; Cycle phase × DayOfWeek) were tested to determine whether environmental timing moderated multiday cycle-related symptom fluctuations. For environmental effects, pairwise comparisons were conducted using Tukey adjustment following the omnibus test.

## Results

### Participant sample

The primary analysis (Model A) included 864 observations from 360 participants. As participants entered the longitudinal study at different timepoints, they completed between one to four assessments (median = 2) with 31.9% completing one assessment, 23.6% two assessments, 16.9% three assessments, and 27.5% four assessments. Model B restricted analyses to participants with at least one observation in both Peak and Trough phases (538 observations; 167 participants), and Model C further restricted to participants with at least two observations in each phase (168 observations; 42 participants). Participant-specific dominant multiday cycle lengths ranged from 6.5 to 58.5 days in the Model A sample. For descriptive and sensitivity analyses, cycle lengths were grouped into four categories: about-weekly (2–12 days), about-fortnightly (13–22 days), about-monthly (23–32 days), and longer cycles (33–60 days). These categories were used to assess whether Peak and Trough observations were drawn from comparable cycle-length distributions.

### Cycle-length characteristics

Cycle length was constant within participants, representing the dominant multiday rhythm selected for each individual across their best segment of heart rate data. Across Peak and Trough observations, represented cycle lengths were similar: Trough observations had a mean cycle length of 26.7 ± 14.3 days, and Peak observations had a mean cycle length of 26.0 ± 14.2 days. At the category level, the distribution of cycle-length categories did not differ significantly between Peak and Trough observations, χ²(3) = 5.26, *p* = 0.154; Fisher’s exact p = 0.154. At the participant level, cycle-length distributions differed significantly by sex, χ²(3) = 83.17, *p* < 2.2 × 10⁻¹⁶; Fisher’s exact p < 2.2 × 10⁻¹⁶, with about-weekly cycles more common among males and about-monthly cycles more common among females. An overview of the cycle length characteristics is presented in Figure 2.

**Figure 2.**
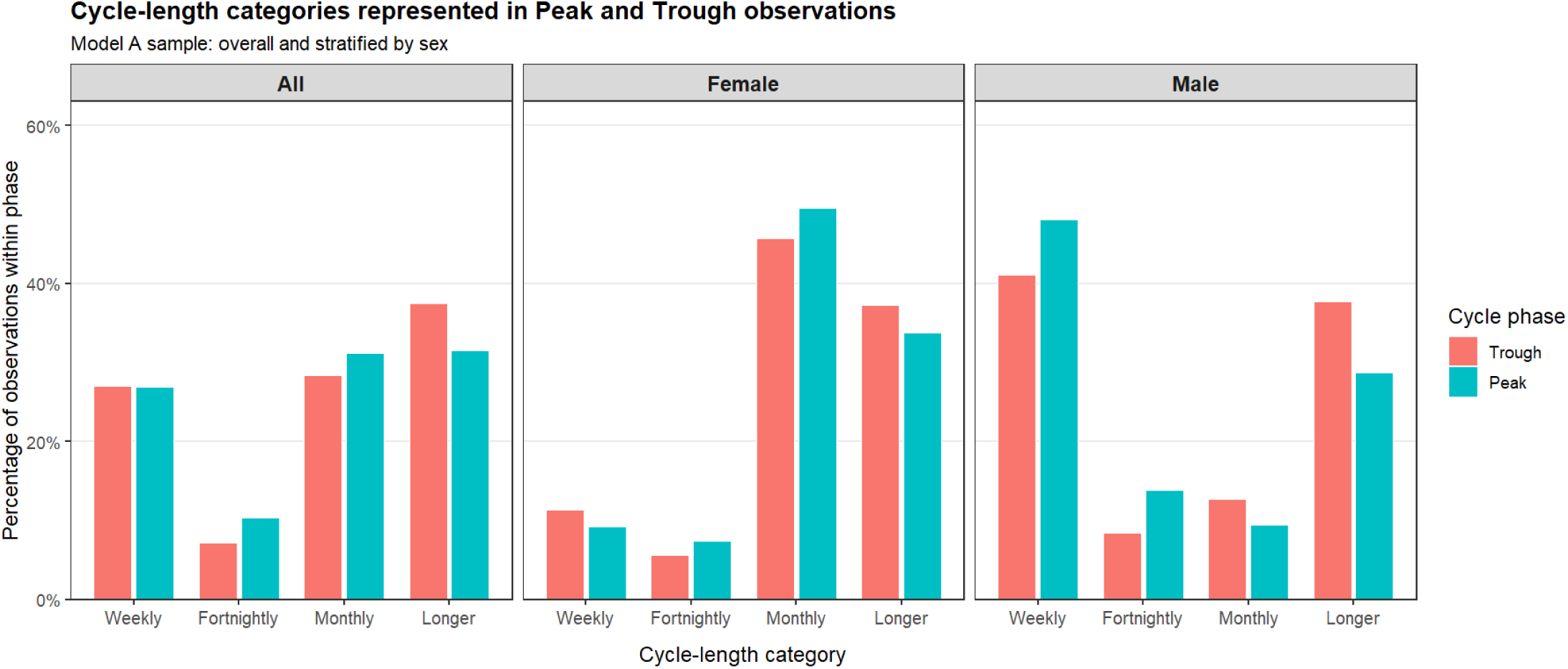
Cycle length characteristics across the aggregated group and when stratified by sex. Cycle lengths were grouped into four categories: Weekly = 2–12 days, Fortnightly = 13–22 days, Monthly = 23–32 days, and Longer = 33–60 days. Overall, Peak and Trough observations were similarly distributed across cycle-length categories (χ²(3) = 5.26, *p* = 0.154; Fisher’s exact *p* = 0.154). Cycle length categories differed by sex, with Weekly cycles more common among males and Monthly cycles more common among females.

### Primary mixed-effects model (Model A)

In the full model, the averaged Cycle Phase effect was not significant, indicating that depressive symptom severity did not differ consistently between Peak and Trough across the full sample. Given our interest in exercise as the primary behavioural moderator, we next examined whether phase-related symptom variation differed by Exercise Level. The Cycle phase × Exercise Level interaction was significant (*F*(1, 712.21) = 5.13, *p* = 0.0238), indicating that Peak–Trough differences depended on exercise level. Planned contrasts showed that within the Low-exercise level, BDI scores were significantly higher during Peak than Trough (Peak − Trough Δ = 1.69, 95% CI [0.11, 3.28], *p* = 0.0366), whereas no Peak–Trough difference was observed in the Higher-exercise level (Δ = −0.43, 95% CI [−1.40, 0.54], p = 0.386). The Type III model-term test also showed an overall Exercise level effect, (*F*(1, 744.22) = 6.62, *p* = 0.010), with higher depressive symptom scores observed in the Low-exercise level overall. Sex also showed a small main effect (*F*(1, 350.62) = 4.53, *p* = 0.034), with females reporting slightly higher BDI scores. Daily mean heart rate was a significant covariate (*F*(1, 466.10) = 14.31, *p* = 0.00018), with higher heart rate associated with greater depressive symptom severity. Together, these results indicate that phase-related symptom variation was conditional on exercise level (see Table 2 and Figure 3).

**Figure 3.**
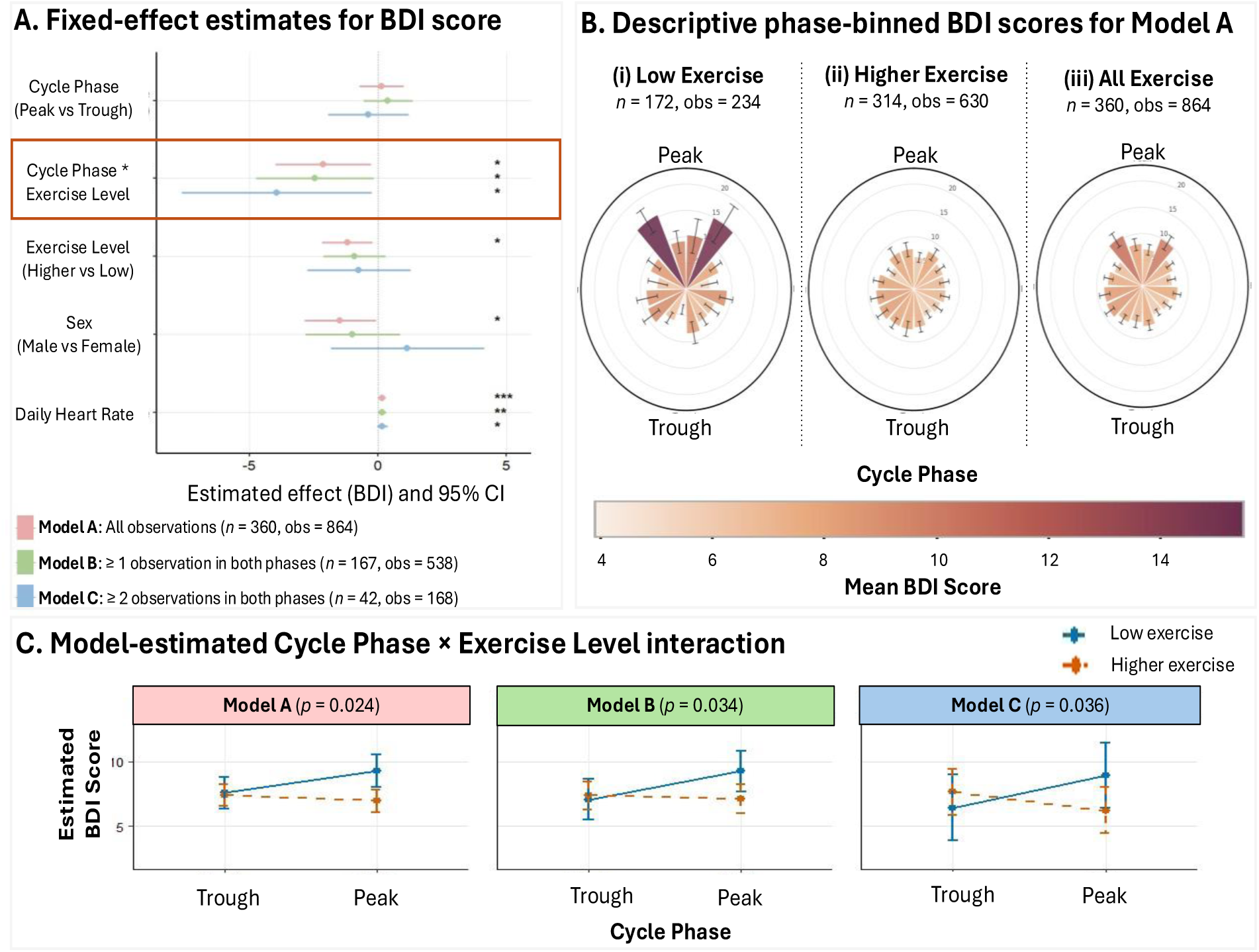
Modelled and descriptive associations between multiday phase, exercise level, and depressive symptom severity. **3A. Fixed-effect estimates from mixed-effects models predicting BDI scores.** Points show model-estimated effects with 95% confidence intervals for the primary and robustness models. For categorical predictors, effects are plotted as contrasts; for Daily Heart Rate, the plotted value is the continuous fixed-effect coefficient. Model A included all eligible observations (*n* = 360 participants, 864 observations), Model B included participants with at least one observation in both Peak and Trough phases (*n* = 167 participants, 538 observations), and Model C included participants with at least two observations in each phase (*n* = 42 participants, 168 observations). All models included a random intercept for participant. Categorical contrasts are shown for Peak versus Trough, the difference in Peak–Trough effects between the higher and low exercise level, higher versus low exercise level, and male versus female. Daily Heart Rate represents the fixed-effect coefficient for centred daily heart rate. Asterisks indicate *p* < 0.05. **3B. Descriptive phase-binned BDI scores for Model A.** Descriptive phase-binned polar plots for Model A show mean BDI score across the reconstructed multiday heart-rate cycle according to low exercise, higher exercise level and BDI score across all observations. Peak is shown at the top and trough at the bottom; bar height and colour indicate mean BDI score within each phase bin. Exercise was self-reported at the survey session, and could change between sessions, therefore participants contributed observations to both exercise levels. These plots are descriptive and do not account for repeated observations within participants. **3C. Model-estimated Cycle Phase x Exercise Level interaction:** Estimated marginal mean BDI scores by cycle phase and exercise level for Models A–C (described above). Error bars show 95% confidence intervals, and interaction p-values are shown above each panel. All models included a participant-level random intercept to account for repeated observations within individuals.

**Table 2.** Model-estimated contrasts from mixed-effects models predicting BDI symptom severity. Estimates are shown in BDI-score units with 95% confidence intervals. Model A included all eligible observations; Model B included participants with at least one observation in both Trough and Peak phases; and Model C included participants with at least two observations in each phase. Cycle Phase estimates represent the model-estimated Peak versus Trough contrast averaged across exercise levels. Exercise Level estimates represent Higher versus Low exercise level. Sex estimates represent Male versus Female. The Cycle Phase × Exercise Level term represents the difference in Peak–Trough contrasts between Higher and Low exercise levels. As cycle-phase effects differed by exercise level, the averaged Cycle Phase contrast should be interpreted cautiously; the key comparisons are the Peak–Trough contrasts within the Low- and Higher-exercise levels reported in the Results text. Daily Heart Rate represents the fixed-effect coefficient per 1-BPM increase in grand-mean-centred daily mean heart rate. All models included a random intercept for participant. Model A was treated as the primary model, whereas Models B and C were sensitivity analyses assessing robustness across increasingly stringent observation requirements. Bold values indicate statistically significant effects (*p* < 0.05).

| Model | Contrast / predictor | $\beta$ | SE | t(df) | 95% CI | p |
| --- | --- | --- | --- | --- | --- | --- |
| **Model A**<br>*n* = 360<br>*Obs* = 864 | Cycle Phase: Peak vs Trough | 0.14 | 0.43 | 0.34 (747.17) | [-0.70, 0.99] | 0.737 |
|  | Cycle Phase × Exercise Level | -2.12 | 0.94 | -2.26 (712.21) | [-3.96, -0.28] | 0.024 |
|  | Exercise Level: Higher vs Low | -1.19 | 0.49 | -2.45 (744.07) | [-2.14, -0.24] | 0.015 |
|  | Sex: Male vs Female | -1.46 | 0.69 | -2.13 (350.62) | [-2.82, -0.11] | 0.034 |
|  | Daily Heart Rate | 0.18 | 0.05 | 3.79 (467.36) | [0.09, 0.27] | <0.001 |
| **Model B**<br>*n* = 167<br>*Obs* = 538 | Cycle Phase: Peak vs Trough | 0.40 | 0.48 | 0.83 (423.69) | [-0.54, 1.33] | 0.408 |
|  | Cycle Phase × Exercise Level | -2.44 | 1.15 | -2.12 (438.77) | [-4.70, -0.18] | 0.034 |
|  | Exercise Level: Higher vs Low | -0.90 | 0.61 | -1.48 (461.67) | [-2.10, 0.29] | 0.139 |
|  | Sex: Male vs Female | -0.98 | 0.93 | -1.06 (167.96) | [-2.81, 0.85] | 0.291 |
|  | Daily Heart Rate | 0.19 | 0.06 | 2.98 (235.06) | [0.06, 0.31] | 0.003 |
| **Model C**<br>*n* = 42<br>*Obs* = 168 | Cycle Phase: Peak vs Trough | -0.36 | 0.78 | -0.46 (133.12) | [-1.90, 1.18] | 0.644 |
|  | Cycle Phase × Exercise Level | -3.92 | 1.86 | -2.11 (139.17) | [-7.59, -0.25] | 0.036 |
|  | Exercise Level: Higher vs Low | -0.73 | 1.01 | -0.73 (152.72) | [-2.72, 1.25] | 0.467 |
|  | Sex: Male vs Female | 1.15 | 1.46 | 0.79 (39.45) | [-1.81, 4.11] | 0.437 |
|  | Daily Heart Rate | 0.18 | 0.09 | 2.02 (50.95) | [0.00, 0.35] | 0.048 |

### Robustness of the primary model as compared to Model B and C

The Cycle phase × Exercise Level interaction was also observed in Model B (*F* (1, 438.77) = 4.50, *p* = 0.0344) and Model C (*F* (1, 139.17) = 4.46, *p* = 0.036). In Model B, higher BDI scores were observed in the Peak relative to Trough phase in the low-exercise level (Δ = 2.21, 95% CI [0.30, 4.12], *p* = 0.0235). In Model C, within-group contrasts did not reach statistical significance (Low exercise: *p* = 0.109), although the direction and magnitude of effects were consistent with Models A and B, supporting the robustness of the model. This pattern suggests that the interaction was retained under increasingly stringent phase-sampling criteria, although within-group contrasts in Model C were less precisely estimated because of the smaller sample. Heart rate remained a significant covariate in Models B and C (*p* = 0.00328; *p* = 0.0499, respectively).

### Sensitivity analysis including participants without significant cycles

Participants with and without significant multiday heart-rate cycles did not differ significantly in BDI scores (Welch’s *t* = 0.30, *p* = 0.763; Wilcoxon *p* = 0.970) or mean daily heart rate (Welch’s *t* = −0.27, *p* = 0.787; Wilcoxon *p* = 0.980). The primary moderation model was then repeated in the expanded eligible sample, without restricting analyses to participants with statistically significant multiday cycles. This model included 955 observations from 413 participants, of which 864 observations were classified as significant-cycle observations. The Cycle Phase × Exercise Level interaction remained significant in the expanded sample, *F*(1, 834.77) = 5.06, *p*=0.0248. This finding was also retained when cycle significance was included as an additional covariate, *F*(1, 781.32) = 6.36, *p*= 0.0119. These sensitivity analyses indicate that the primary interaction effect was not solely driven by restricting the analysis to participants exceeding the permutation-based significance threshold.

### Environmental and calendar effects

(see Figure 4): Season and Day of Week were examined to explore environmental and calendar effects on symptom variability across the study period. Only Winter, Spring, and Summer were represented, as no BDI assessments occurred during Autumn. Depressive symptoms differed by season (*F*(2, 652.49) = 7.15, *p* = 0.00085), with Tukey-adjusted pairwise comparisons indicating higher symptoms in Winter relative to Spring (Δ = 1.636, 95% CI [0.562, 2.711], *p* = 0.0011), whereas Winter did not differ significantly from Summer (Δ = 1.005, 95% CI [−0.180, 2.189], *p* = 0.1148) and Spring did not differ from Summer (Δ = −0.632, 95% CI [−1.754, 0.491], *p* = 0.3833). Day of Week showed no significant association with depressive symptoms (*F*(6, 727.91) = 0.726, *p* = 0.629).

**Figure 4.**
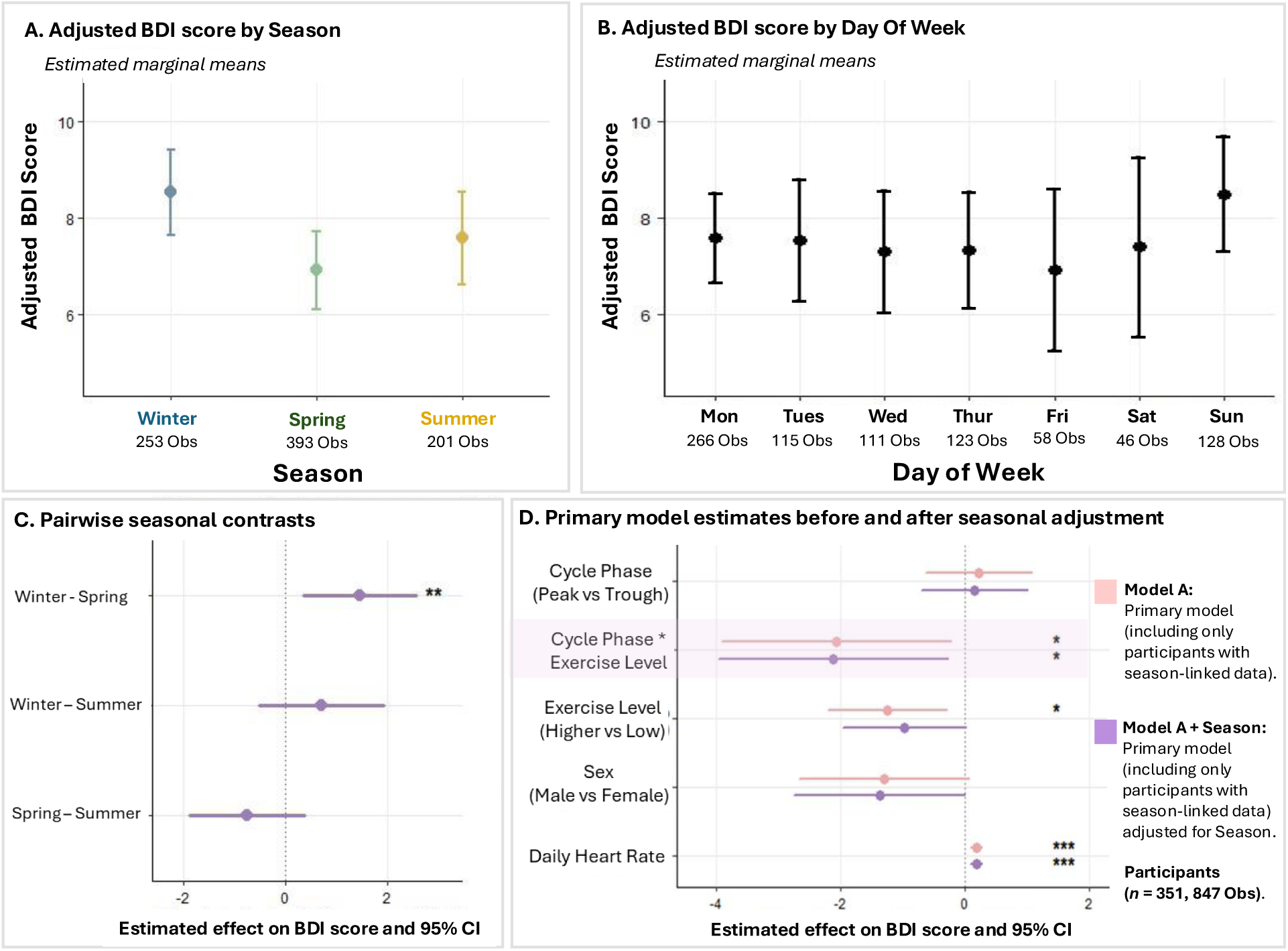
Environmental timing effects and seasonal robustness of primary multiday-cycle model. This figure presents estimated marginal means and 95% confidence intervals for BDI score by **4A. Season** and **4B. Day of Week** from separate random-intercept models examining the association between environmental/calendar timing and BDI symptom severity. These models contextualise symptom variability across the study period before adding environmental covariates to the primary model. Season showed a significant association with BDI, no significant association was observed with Day of Week. **4C Pairwise seasonal contrasts.** Shows Tukey-adjusted pairwise season contrasts from the primary mixed-effects model additionally adjusted for Season. Winter was associated with higher estimated BDI scores than Spring, whereas Winter did not differ significantly from Summer, and Spring did not differ significantly from Summer. **4D. Primary model estimates before and after seasonal adjustment.** Presents model-estimated contrasts and 95% confidence intervals for the primary model (Model A, pink lines) and then Model A when adjusted for Season (purple lines). Season was included as a covariate in the adjusted model, with Season contrasts shown in Panel C. The Cycle Phase × Exercise Level interaction remained significant after including Season, indicating that the primary phase-linked effect was not explained by seasonal differences in overall depressive symptom severity. Both models included participant-level random intercepts.

Season was then added as a covariate to the primary mixed-effects model (Model A). Including Season improved model fit (likelihood ratio test χ²(2) = 10.47, *p* = 0.00533), with Season showing a significant main effect in the adjusted model (*F*(2, 625.79) = 5.22, *p* = 0.00564). Tukey-adjusted pairwise comparisons in the adjusted model indicated higher depressive symptoms in Winter relative to Spring (Δ = 1.49, 95% CI [0.40, 2.58], *p* = 0.0038), with no significant differences between Winter and Summer (Δ = 0.78, 95% CI [−0.44, 2.00], *p* = 0.2869) or Spring and Summer (Δ = −0.71, 95% CI [−1.83, 0.41], *p* = 0.2972). After adjusting for Season, the Cycle Phase × Exercise Level interaction remained significant (*F*(1, 710.68) = 5.39, *p* = 0.0205). Adding a Season × Cycle Phase interaction did not improve model fit (χ²(2) = 0.97, *p* = 0.615), indicating that seasonal context contributes additively to overall symptom severity but did not modify the multiday cycle-related effects.

Day of Week was not associated with depressive symptoms in Model A, and the Cycle Phase × Exercise Level interaction remained significant after adjusting for DayOfWeek (*F*(1, 710.45) = 5.61, *p* = 0.01810). Adding Day of Week did not improve model fit (χ²(6) = 4.825, *p* = 0.5665), and there was no evidence of DayOfWeek × Cycle phase moderation (χ²(6) = 9.285, *p* = 0.158).

## Discussion

The present study introduces a wearable-informed framework for testing whether depressive symptoms fluctuate with person-specific infradian rhythms. Across three increasingly stringent sampling models, depressive symptoms were elevated near the peak of an individual’s cycle among participants reporting lower exercise. This association was exercise-dependent; peak-trough differences were evident during low exercise, whereas no comparable difference was observed during higher exercise. Established correlates of depressive symptoms, including daily heart rate, sex, exercise level, and seasonality, behaved largely as expected, supporting the biological plausibility of the modelling framework. The Cycle Phase X Exercise Level interaction remained evident after accounting for these covariates, while additional candidate features, including sleep metrics, day of week, and multiday cycle length, did not improve model fit. Together, these findings indicate that person-specific infradian phase may index a temporal “when” component of depressive symptom vulnerability that extends beyond daily autonomic arousal, seasonal context, or demographic factors.

### Multiday cycle phase as a marker of depressive mood symptoms

Despite evidence for infradian rhythms in autonomic, endocrine, and neurological systems (18,19,28,49), longitudinal studies of depressive symptoms rarely test whether symptom fluctuations show person-specific rhythmicity over timescales of weeks to months. Here, we leveraged continuous wearable-derived signals to estimate individualised multiday phase and linked this physiological timing marker to repeated depressive symptom assessments. Depressive symptoms were higher near cycle peak than trough during periods of lower exercise, suggesting that longer-timescale symptom shifts may reflect structured temporal organisation rather than stochastic variation alone. This interpretation is consistent with earlier psychiatric studies that reported stable person-specific periodicities in mood over days to months in both affective-disorder samples (6–10) and healthy cohorts (3–5,38). This interpretation also aligns with earlier chronobiological evidence that mood and affective symptoms can show non-24-hour temporal structure, including multiday variation in positive and negative affect among healthy students (35), 48-hour mood and sleep–wake cycling in affective illness (11,36,60), and infradian cardiovascular components (blood pressure and heart rate) during depression (37). Recent circuit-level work further supports the biological plausibility of infradian affective-relevant rhythms, showing that mesolimbic dopamine neurons (altered in depressive disorders) can drive multiday rhythms in sleep–wake timing, locomotor activity, and heightened activity states in mice (61). When combined, these studies suggest that multiday mood variability should not be treated solely as noise but may partly reflect clinically meaningful (6), structured within-person dynamics (7). The present study revisited this question by using continuous wearable physiology, allowing multiday phase to be estimated passively and linked to symptom timing in a scalable manner. Our results also align with work in other clinical domains showing that multiday peak phase can coincide with heightened vulnerability markers, including increased seizure likelihood (62) and elevated stress-hormone levels (28). Together, these findings support the possibility that multiday peak phase indexes a broader physiological state of increased vulnerability, although causal mechanisms require further clarification.

After the 1990s, research into individual-level rhythmic structure declined significantly, and longitudinal studies focused more heavily on calendar-defined windows, such as seasonality or menstrual-cycle timing. However, these approaches typically rely on group-level comparisons or predefined calendar-based timepoints and therefore have limited capacity to capture person-specific symptom rhythms that vary across individuals. In the present study, depressive symptoms were higher in winter relative to spring, consistent with prior work on seasonality (43). However, while seasonal context contributed independently to symptom levels, adjusting for season did not attenuate the Cycle Phase X Exercise Level association. These findings suggest that depressive symptom vulnerability may be shaped by multiple temporal scales: seasonal context may contribute to overall symptom burden, whereas person-specific infradian phase indexes the timing of within-person symptom fluctuation.

The broader pattern of associations identified in this study supports the biological plausibility of the modelling framework. In individuals with a significant infradian rhythm, higher daily heart rate was associated with greater depressive symptom severity (63,64), consistent with autonomic dysregulation models of depression (50,53), and prior evidence linking elevated resting heart rate and reduced heart rate variability with depressive symptoms (51,65,66). Importantly, the phase-linked pattern remained evident after adjusting for daily heart rate, suggesting that personalised multiday timing indexes information beyond day-level autonomic arousal. Lower exercise was associated with higher depressive symptom severity and moderated the phase-related effect, with peak–trough differences observed primarily during lower-exercise periods. This aligns with evidence that physical activity is associated with lower depressive symptom burden (58,59), while suggesting that exercise may also influence the expression of mood variation across physiologically defined multiday phase. In this healthy young cohort, lower exercise may have marked periods of greater symptom variability, making phase-related differences more detectable. Conversely, higher exercise may have been associated with attenuated phase-linked symptom differences, consistent with the proposed protective role of physical activity against depressive symptoms (58). The association between female sex and higher depressive symptom severity was also consistent with population-level patterns (67,68). Taken together, these findings indicate that, consistent with past studies, seasonality, daily heart rate, exercise level, and sex contribute to overall depressive symptom burden, while suggesting that person-specific infradian phase may capture an additional source of variation.

In psychiatric research, symptom fluctuation is a core feature of many disorders (30,42), ranging from mood lability in bipolar disorder to episodic symptom exacerbations across affective disorders. Although such fluctuations are often attributed to environmental drivers, many studies are not designed with the sampling density and follow-up needed to map person-specific symptom trajectories. Building on the present results, we propose that future longitudinal designs consider incorporating multiday rhythms, captured non-invasively using continuous monitoring together with more frequent symptom sampling, to map individual mood trajectories. Within-person symptom shifts should not be dismissed as measurement noise; rather, our findings suggest that a component of symptom variability may reflect longer-timescale temporal organisation (11,35–37,60). This point echoes the conclusions of the earlier mood-variability studies, which argued that the timing of symptom change within an individual may itself be clinically informative. If symptom levels fluctuate over multiday rhythms, this has implications for mood research, whereby single-session questionnaires or poorly timed assessments may introduce additional variance and obscure brain–symptom associations. Rhythm-aware sampling, which aligns symptom assessments (and potentially neuroimaging sessions) to person-specific phase, may help us understand the variance and improve reproducibility across studies.

### Personalised multiday cycles and digital technology

Digital phenotyping has created new opportunities to monitor mental health outside the clinic (69–72) and track symptom change (73,74). Many current approaches focus on classification models that distinguish between depressed and non-depressed groups (75), quantify depression severity (70) or predict short-term daily risk (76,77), using passive features such as movement, light exposure, sleep, and heart rate variability. Given rapid advances in wearable technologies and the increasing availability of continuous monitoring data, many large-scale longitudinal studies are now collecting passive data, including exercise, heart rate, sleep, and step count, alongside self-report mood measures (78,79), with the aim of predicting depressive symptoms using digital phenotyping approaches. This emerging wave of longitudinal data collection provides an important foundation for passive mental-health monitoring, but it does not yet establish whether symptom vulnerability follows recurring, person-specific temporal rhythms. The present study addresses this gap by applying cycle-based modelling to test whether depressive symptom severity varies according to an individual’s own multiday physiological phase. Rather than focusing only on symptom presence or severity on a given day, cycle-based modelling seeks to identify recurring windows of heightened vulnerability within individuals. This is important because personalised models in psychiatry have often focused on symptom profiles, treatment selection, or dosage considerations, with comparatively less attention to the timing of symptom change. Additionally, while developing predictive models was beyond the scope of the present study, our initial findings lay the groundwork for testing whether infradian phase can be leveraged to improve future digital models of depressive symptom escalation

### Limitations

Several limitations should be acknowledged. First, the sample comprised healthy college-aged adults, which may limit the generalizability of the findings to clinical populations or other age groups. We intentionally studied a healthy sample to validate the cycle-based model and establish a normative baseline without the additional confounding influences of antidepressant medication, acute illness, or more chronic symptom burden; the next step is to apply this framework in a clinical cohort. Second, the observational design limits causal inference. While multiday cycle phase was associated with symptom fluctuations, the present study cannot determine whether physiological rhythms influence mood, mood-related changes alter autonomic physiology, or both reflect shared upstream processes such as stress, sleep, endocrine state, or social rhythms. However, the ability to non-invasively monitor individuals in a naturalistic setting and identify these patterns using wearable devices for continuous monitoring remains a notable strength of the study. Third, symptom severity was assessed using self-report measures (BDI), a widely validated self-report measure. Depressive symptoms were assessed intermittently rather than daily, therefore, the present study cannot establish the full waveform, temporal stability, or periodicity of symptom fluctuations. Instead, it tests whether symptom severity differs by physiologically defined multiday phase. Future studies should combine validated symptom scales with higher-frequency ecological momentary assessment to determine whether multiday phase relates to shorter-term mood dynamics. Fourth, cycle detection depends on sufficient data continuity and signal quality. Variability in wearable adherence, missingness, and physiological noise may affect phase estimation. In addition, the permutation-based approach used to identify significant multiday cycles removes both rhythmic structure and autocorrelation and may therefore provide a liberal test of rhythmicity. Although sensitivity analyses showed that the primary Cycle Phase × Exercise Level interaction persisted when participants without statistically significant cycles were included, future work should use autocorrelation-preserving surrogate data to test whether detected multiday peaks exceed those expected from serially dependent physiological noise. Therefore, while it was beyond the scope of the current study, as wearable-derived multiday rhythm detection grows as a promising technique, it would be worthwhile for future studies to optimise these pipelines and systematically compare alternative approaches for identifying statistically significant individual-level cycles. However, despite this, prior work using long-term heart rate (17,18) data suggests that multiday rhythms can be reliably extracted from naturalistic recordings when sufficient data are available. Finally, the present model did not account for all environmental and biological contextual factors that may influence mood or physiology, including academic stress, social rhythms, medication, illness or menstrual phase, hormonal status, and contraceptive use. Academic stress is particularly relevant in college-aged cohorts because assessment periods, deadlines, and semester timing may influence both mood and physiology. Menstrual phase could not be modelled directly, which is an important limitation. However, cycle length did not improve model fit, and peak and trough observations were drawn from comparable cycle-length distributions, suggesting that the observed association was unlikely to be explained solely by a monthly menstrual rhythm. This does not rule out menstrual or hormonal influences, and future studies should directly incorporate additional contextual factors such as menstrual phase, hormonal status, and contraceptive use where relevant. Sleep measures were also not significantly associated with depressive symptoms in this healthy young cohort, which may reflect restricted sample variability and should not be interpreted as contradicting established sleep-mood relationships. Further research is warranted to explore the relationship between sleep, depressive symptoms, and multiday phase in a more heterogenous sample.

## Conclusion

This study provides evidence that depressive symptoms fluctuate over long-term physiological rhythms derived from heart rate. By estimating multiday cycle phase from continuous wearable physiology, this framework enables structured mood periodicity to be revisited using scalable, non-invasive methods. These findings lay the groundwork for data-driven, personalised approaches to understanding mood dynamics and motivate future studies in clinical populations to test whether cycle-based models can track patterns of symptom vulnerability. More broadly, this approach advances digital mental health by enabling low-burden remote monitoring of patterns of symptom vulnerability. Ultimately, such models could help clinicians and patients move toward personalised monitoring frameworks that identify periods of elevated vulnerability, with the longer-term goal of supporting earlier and better-timed intervention strategies.

## Data availability

The anonymised data collected are available as open data via the University of Notre Dame’s (USA) NetHealth online repository: https://sites.nd.edu/nethealth/

## Acknowledgements

P.J.K acknowledges funding by National Health and Medical Research Council (APP2033596) and the Australian Research Council (DP240102899). These funders played no role in study design, data collection, analysis and interpretation of data, or the writing of this manuscript.

